# Design and Retrospective Cohort Validation of an Algorithm for Surgical Site Infection Surveillance after Hip Replacement Surgery Using Natural Language Processing and Extreme Gradient Boosting

**DOI:** 10.1101/2023.01.17.23284669

**Authors:** Alvaro Flores-Balado, Carlos Castresana Méndez, Antonio Herrero González, Raúl Mesón Gutierrez, Javier Arcos, María Dolores Martín-Ríos, the Surgical Site Infection Surveillance Group

## Abstract

**Background:** Surgical site infection (SSI) surveillance is a labor-intensive endeavor. We present the design and retrospective cohort validation of a multivariable algorithm to screen for SSI in patients undergoing hip replacement surgery, permitting real-time case detection, and reducing the number of clinical records to be reviewed manually.

**Methods:** We designed a multivariable algorithm using extreme gradient-boosting (XGBoost) to screen for SSI in patients undergoing hip replacement surgery. The development and validation cohort included all healthcare episodes dating from the initial hospital admission for surgery until 90 days after joint replacement (n=19661) between January 2014 and December 2021 from four hospitals in Madrid, Spain. A NLP pipeline was implemented to obtain text variables from free-text fields of the EHR. Clinical, prescription, microbiology, and laboratory variables were also collected. Episodes were split randomly into training (70%) and testing (30%) datasets. Hyperparameters were adjusted to penalize false negatives.

**Findings:** The presence of positive microbiological cultures, the text variable “infection”, and the prescription of clindamycin were strong markers of SSI. Statistical analysis of the final model indicated a high sensitivity (99·18%) and specificity (91·01%) with an F1-score of 0·32, AUC of 0·989 and accuracy of 91·27%. The model correctly classified 5129 episodes as negative for SSI, with only one case of SSI escaping detection (NPV 99·98%).

**Interpretation:** We conclude that the combination of NLP and extreme gradient boosting is a sensitive tool for SSI surveillance in hip replacement surgery, permitting real-time, semi-automatic surveillance. In practice, our results translate as an 88·95% reduction in the total volume of clinical records to be reviewed manually. To the best of our knowledge, this is the first time that an algorithm incorporating data from multiple sources using NLP and extreme gradient boosting has been developed for orthopedic SSI surveillance.

**Funding:** This study did not receive any funding.

## Introduction

Surgical site infection (SSI) surveillance is a recurring priority in healthcare policy. SSIs account for up to 20% of nosocomial infections^1^, increasing mortality and length of hospital stay, as well as the need for unplanned hospital admissions, new surgical interventions, and ICU readmissions ^2,3^. Although constant SSI surveillance is a fundamental pillar of infection prevention and outbreak control, it remains a labor-intensive endeavor requiring the dedication of valuable time and resources, with up to 45% of infection prevention staff time dedicated to surveillance^4^. While both in-house and commercial infection control surveillance software have been developed, most hospital SSI surveillance programs still rely heavily on the manual review of medical records^5,6^, leading to workload-related challenges for universal surveillance and case detection. In this setting, real-time screening for SSI is far from plausible.

The increasing demand for joint replacement surgery^7^, the emergence of multi-drug resistant pathogens^8^, and the devastating consequences of prosthetic joint infection^9^ lead to an urgent need for SSI surveillance optimization in the field of orthopedics. With the widespread implementation of electronic health records (EHRs) as the current standard of clinical data management, several strategies for orthopedic SSI surveillance using natural language processing (NLP) and logistic regression models have been described and implemented in clinical practice^3,10,11^. However, the combination of NLP with other machine learning techniques has yet to be explored.

We present the design and retrospective cohort validation of a multivariable algorithm to screen for surgical site infection in patients undergoing hip replacement surgery as part of a semiautomated approach to SSI surveillance, permitting real-time case detection and reducing specialist workload.

## Methods

### Study design

We designed a multivariable algorithm to screen for SSI in patients undergoing hip replacement surgery. Our algorithm development and validation cohort included all patients undergoing hip replacement surgery (n = 6741, corresponding to 7444 surgeries) between January 2014 and December 2021 from four hospitals (Fundación Jiménez Díaz University Hospital, Infanta Elena University Hospital, Rey Juan Carlos University Hospital, and Central Villalba University Hospital) belonging to the Quironsalud 4-H Network, Madrid, Spain.

All healthcare episodes (hospital admissions, emergency department visits, and outpatient clinic appointments) dating from the initial hospital admission for surgery until 90 days after joint replacement were included as separate episodes in our dataset (n=19661). Text variables were obtained for each episode from the free-text fields of the EHR using NLP. Clinical, prescription, microbiology, and laboratory variables were collected from multiple datasources for each episode.

Our objective was to predict the presence of SSI using artificial intelligence (SSS-ai). We defined the gold standard as the diagnosis of SSI documented by clinical experts (SSI-gs). For development and validation, all registers were labelled as positive or negative for SSI-gs following the definitions of the European Centre for Disease Control’s HAI-NET SSI protocol, version 2·2^12^. Both superficial and deep incisional infections were included. 54·5% of surgical interventions (n=4057) had already been screened for SSI by members of the preventive medicine department as part of the Network’s routine SSI surveillance program, while the remaining 3387 were retrospectively screened and labelled by members of the infection control department for the purpose of this study.

A literature review was carried out to search for relevant features for algorithm design and training. The initial results of the review were presented to members of the preventive medicine and orthopedic surgery departments, with the final list of 35 features selected by consensus (table 1).

**Table 1:**
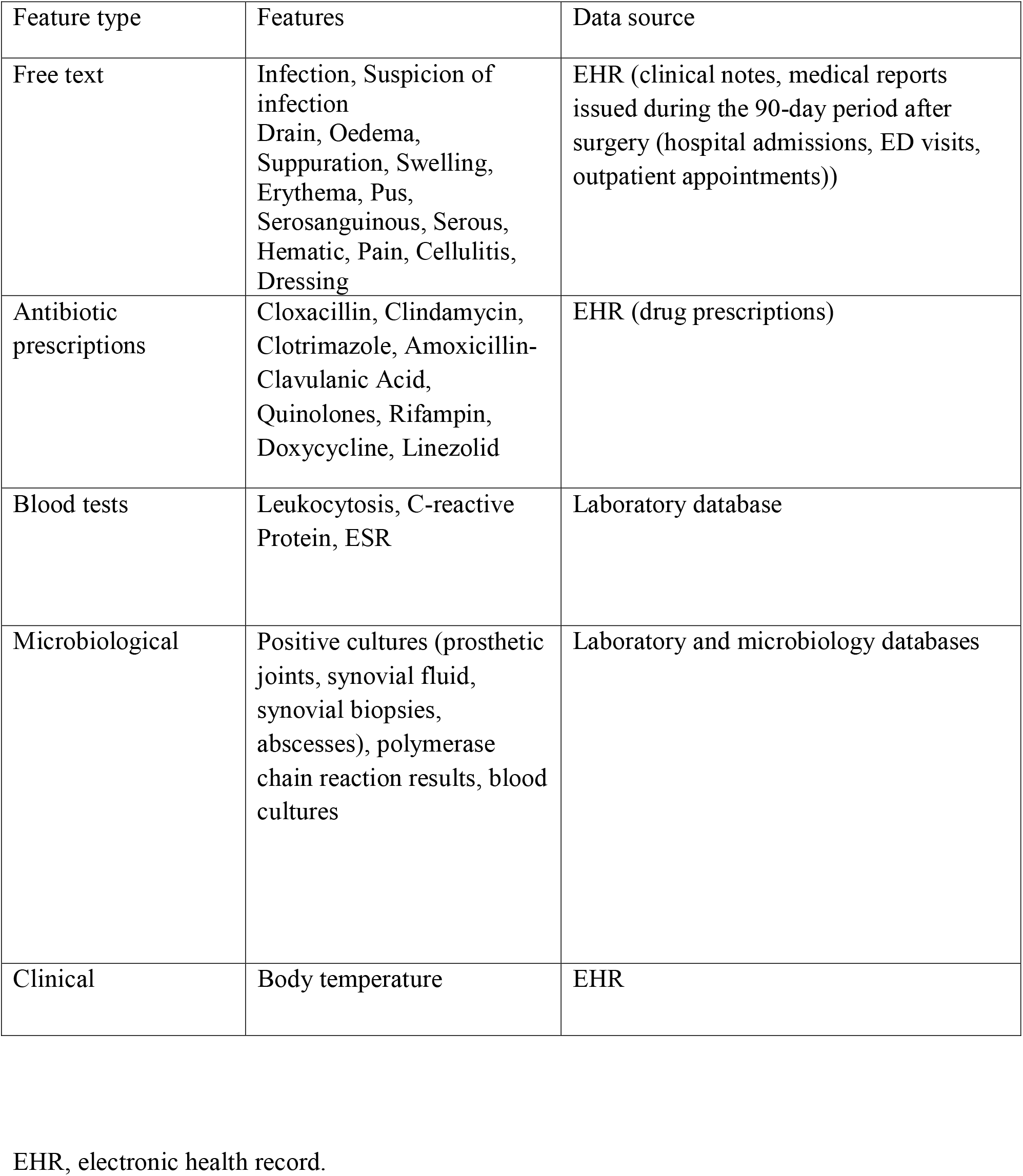
Features used in model development

| Feature type | Features | Data source |
| --- | --- | --- |
| Free text | Infection, Suspicion of infection<br>Drain, Oedema, Suppuration, Swelling, Erythema, Pus, Serosanguinous, Serous, Hematic, Pain, Cellulitis, Dressing | EHR (clinical notes, medical reports issued during the 90-day period after surgery (hospital admissions, ED visits, outpatient appointments)) |
| Antibiotic prescriptions | Cloxacillin, Clindamycin, Clotrimazole, Amoxicillin-Clavulanic Acid, Quinolones, Rifampin, Doxycycline, Linezolid | EHR (drug prescriptions) |
| Blood tests | Leukocytosis, C-reactive Protein, ESR | Laboratory database |
| Microbiological | Positive cultures (prosthetic joints, synovial fluid, synovial biopsies, abscesses), polymerase chain reaction results, blood cultures | Laboratory and microbiology databases |
| Clinical | Body temperature | EHR |

EHR, electronic health record.

Our research protocol conformed to the Declaration of Helsinki and received approval from the institutional ethics committee. TRIPOD standards for development and validation reporting were followed.

### Data preprocessing

#### Data extraction pipeline

In order to conduct real-time data extraction and apply NLP techniques without affecting the network EHR’s (CASIOPEA2®, Inetum) system performance, we replicated the EHR database (Microsoft SQL version 10·50), including information from clinical notes, laboratory values, microbiology results, and drug prescriptions, optimizing extraction speed with temporal tables, cursors, and groupings. An automatic task was programmed to refresh the database every hour.

#### Natural language processing and feature engineering

The inclusion of information from clinical notes posed particular challenges, as the CASIOPEA2 EHR uses free-text fields, leading to unstructured data featuring non-standard abbreviations, spelling mistakes, and frequent synonyms. A NLP pipeline (Pandas, NLTK) was implemented with the objective of identifying target variables and reducing model complexity by transforming them into binary variables (true/false). Accent cleaning and lower-case transformation was carried out. Posteriorly, stop words were removed, screening for relevant abbreviations and modifiers. Sentence splitting and tokenization were carried out and named-entity recognition was used for variable identification.

High cardinality variables, such as hospital admission dates, were eliminated for algorithm development. Qualitative data, such as drug prescriptions, were assigned a classification code to ensure the correct identification of all variables. Laboratory values posed a particular challenge, as several observations per episode were often available. To reduce complexity, minimum, maximum, and median values were calculated for each episode and included in the dataset.

### Model design and testing

Episodes were split randomly into training (70%) and testing (30%) datasets. We trained a binary classification model using extreme gradient-boosting (XGBoost)^13^, which combines decision trees with an adaptative gradient, to predict SSI (SSI-ai) using a set of 35 features selected a priori by members of the preventive and orthopedic departments after an extensive literature review. XGBoost was chosen for algorithm development because of its robust performance in the presence of an elevated number of null values.

As our primary objective was to screen for patients with SSI, hyperparameters were adjusted to improve recall (TP/TP+FN). Given the unbalanced dataset, with a 2·85% overall prevalence of SSI-gs and a 2·26% prevalence of SSI-gs in the development cohort, we applied sample weights to penalize false negatives. Weights were selected as proportionate to the difference in volume between targets (SSI-gs negative: SSI-gs positive, 1:44).

Feature importance was assessed using SHAP values to identify the predictors’ contributions to the final prediction (figure 1), with the 10 most relevant predictors being selected for the final model (figure 2).

**Figure 1:**
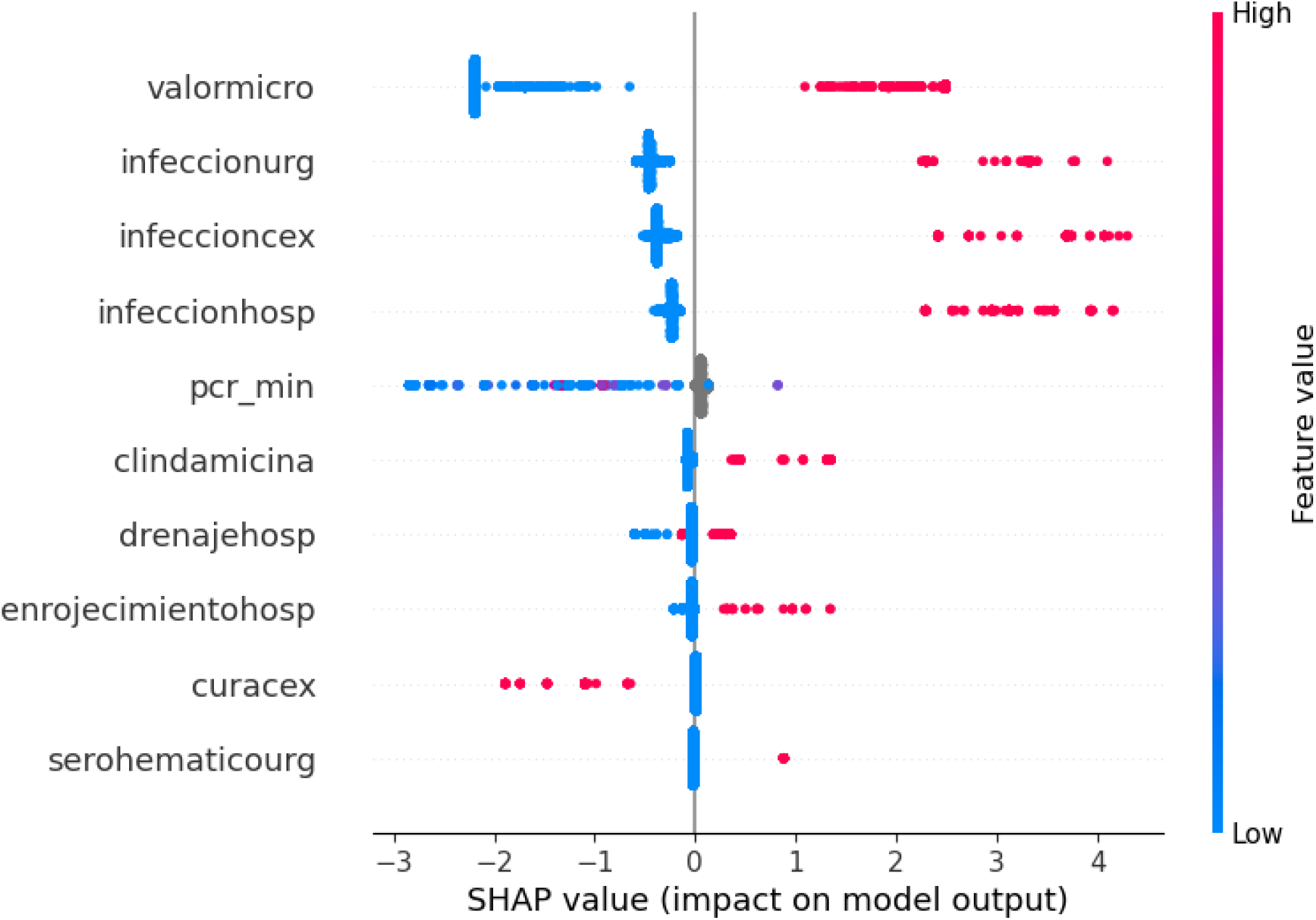
SHAP values for features included in the final model valormicro, presence of a positive microbiological culture; infeccionurg, detection of the free-text entity “infection” in emergency department reports; infeccioncex, detection of the entity “infection” in outpatient appointment reports; infeccionhosp, detection of the entity “infection” in hospital admission or discharge reports; pcr_min, elevated C-reactive protein levels; clindamicina, prescription of clindamycin; drenajehosp, detection of the free-text entity “drain” in hospital admission or discharge reports; enrojecimientohosp, detection of the free-text entity “erythema” in hospital admission or discharge reports; curacex, detection of the free-text entity “dressing” in outpatient appointment reports; serohematicourg, detection of the free-text entity “serosanguinous” in emergency department reports.

**Figure 2:**
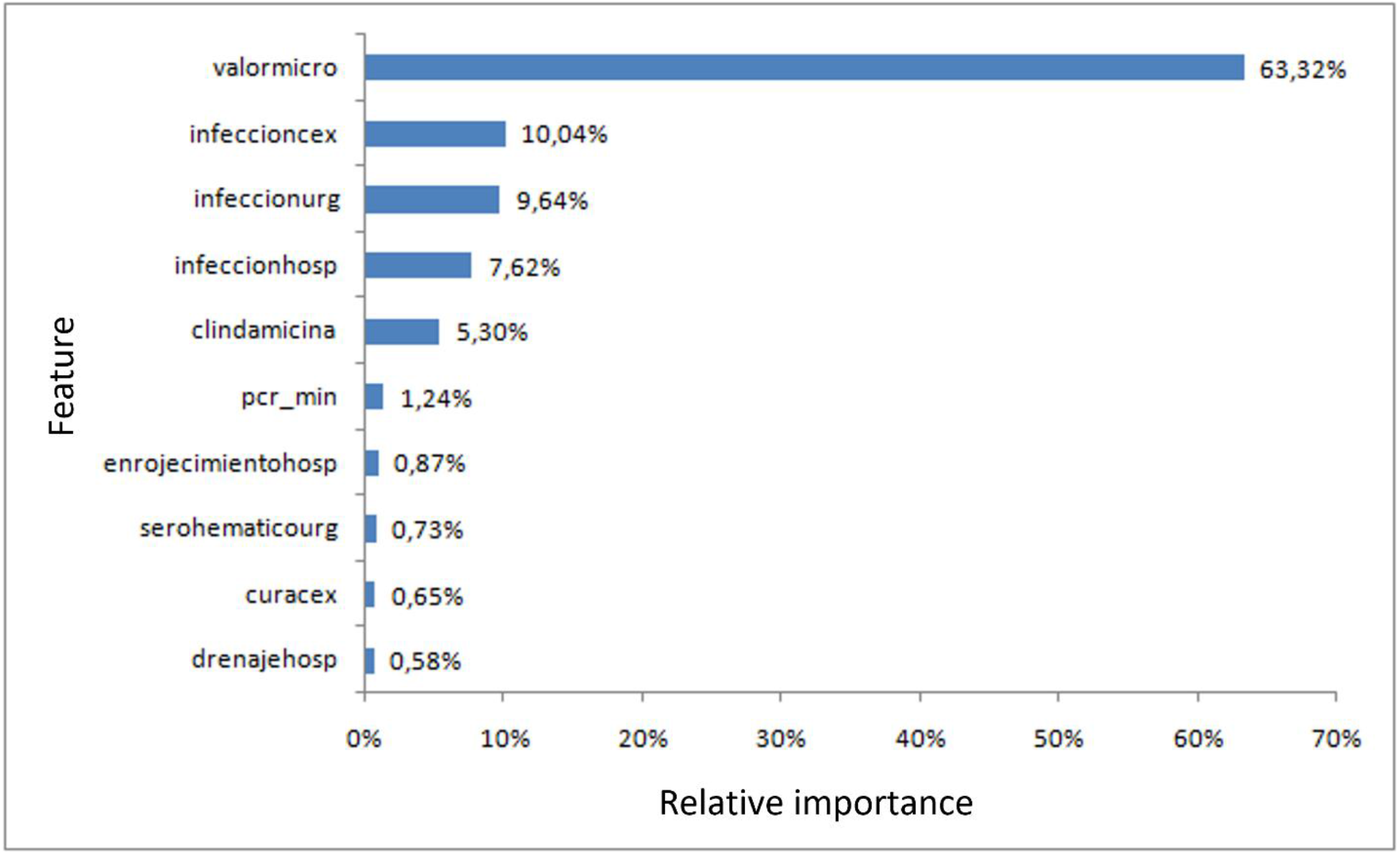
Feature importance valormicro, presence of a positive microbiological culture; infeccionurg, detection of the free-text entity “infection” in emergency department reports; infeccioncex, detection of the entity “infection” in outpatient appointment reports; infeccionhosp, detection of the entity “infection” in hospital admission or discharge reports; pcr_min, elevated C-reactive protein levels; clindamicina, prescription of clindamycin; drenajehosp, detection of the free-text entity “drain” in hospital admission or discharge reports; enrojecimientohosp, detection of the free-text entity “erythema” in hospital admission or discharge reports; curacex, detection of the free-text entity “dressing” in outpatient appointment reports; serohematicourg, detection of the free-text entity “serosanguinous” in emergency department reports.

### Statistical analysis

Model performance was evaluated through analysis of sensitivity, specificity, accuracy, positive predictive value (PPV), and negative predictive value (NPV) using Epidata version 4·2, and the area under the receiver operating characteristic curve (AUC), calculated using Scikit-Learn 1·2·0 (Python 3·9). Categorical variables were compared using a chi-squared test, and p values were set at <0·05 for statistical significance.

## Results

19661 episodes (hospital admissions, emergency department visits, or outpatient clinic appointments during the 90 days following surgery) corresponding to 7444 hip replacement surgeries were included in our dataset, of which 547 (2·85%) were identified as presenting surgical site infection upon manual review of clinical records by members of the preventive medicine department (SSI-gs). 5751 episodes were included in the external validation cohort, with 121 episodes identified as SSI-gs (2·12%).

After model iteration, 10 features were identified as most relevant for the detection of SSI (figure 2), of which the presence of a positive microbiological culture presented the highest relative feature importance (63·32%), followed by the detection of the entity “infection” in outpatient appointment reports (10·04%), emergency department reports (9·64%) or hospital admission and discharge reports (7·62%). The prescription of clindamycin during the 90 days following surgery and elevated C-reactive protein values were the next most relevant features (with a relative feature importance of 5·30% and 1·24%, respectively), while the detection of the entities “erythema” in hospital admission reports (0·87%), “serosanguinous” in emergency department reports (0·73%), “dressing” in outpatient appointment reports (0·65%), and “drain” (0·58%) in hospital admission reports completed the list of features included in the final model. Table 2 depicts the distribution of the five most relevant features between SSI-sg positive and negative subgroups.

**Table 2:** Distribution of most relevant features according to presence of surgical site infection

| Feature | No surgical site infection<br>(SSI-gs negative)<br>N (%) | Surgical site infection<br>(SSI-gs positive)<br>N (%) | p-value |
| --- | --- | --- | --- |
| Positive microbiological culture | 1651 (22.73%) | 173 (95.58%) | <0.0001 |
| Infection_out | 181 (2.49%) | 59 (32.60%) | <0.0001 |
| Infection_ED | 287 (3.95%) | 98 (54.14%) | <0.0001 |
| Infection_hosp | 103 (1.42%) | 93 (51.38%) | <0.0001 |
| Clindamycin | 63 (0.87%) | 30 (16.57%) | <0.0001 |

Infection\_out, presence of the free-text entity “infection” in an outpatient appointment report. Infection\_ED, presence of the free-text entity “infection” in an emergency department report. Infection\_hosp, presence of the free-text entity “infection” in a hospital admission/discharge report.

As the objective of our tool, AI-HPRO, was to screen for SSI and given the unbalanced dataset (2·54% SSI-gs), we applied sample weights to penalize false negatives. Statistical analysis of the final model indicated a high sensitivity (99·18%) and specificity (91·01%) with an F1-score of 0·32, AUC of 0·989 and accuracy of 91·27%. The model correctly classified 5129 episodes as negative for SSI-gs, with only one case of SSI-gs escaping detection (NPV 99·98%). 622 episodes were classified as SSI-ai, with 121 labelled as SSI-gs by members of the preventive medicine department (PPV 19·45%). In practice, these results translate as an 88·95% reduction in the total volume of clinical records to be reviewed manually.

The AI-HPRO screening algorithm is now fully incorporated into the preventive medicine department’s workflow. Where operational aspects are concerned, a user dashboard has been developed to enable members of the preventive medicine department to run the model on-demand, permitting real-time SSI surveillance. The data capture pipeline is fully automated, with a function programmed to refresh the database automatically on an hourly basis. Episodes flagged as positive for SSI-ai by the algorithm are validated manually by members of the preventive medicine team, who either verify the diagnosis of SSI or classify the registers as false negatives. Validation is fed back into the database, and the algorithm retrains using the new inputs, thus ensuring continuous improvement.

## Discussion

Our study presents a novel tool, AI-HPRO, which combines NLP techniques and a multivariable prediction algorithm to screen for SSI in patients undergoing hip replacement surgery, potentially reducing surveillance workload by over 88%. SSI surveillance through the manual review of clinical records is a time-intensive, resource-consuming task. Due to the characteristics of the data generated for each episode of SSI (rapid generation, high-volume, and variability), artificial intelligence is a valuable ally for specialists in preventive medicine^14^. Our semi-automated approach, in which the vast majority of the surveillance burden is handled by artificial intelligence, leaves members of the infection prevention department with the task of ruling out SSI after hip replacement surgery in a reduced subset of possible cases. Surveillance volume reduction and the implementation of an automatic data capture pipeline, which is refreshed on an hourly basis, opens the door to the possibility of real-time SSI surveillance in orthopedics, increasing the efficacy of different preventive strategies and providing new opportunities for outbreak prevention.

A systematic review published in 2020^15^ identified 29 electronic SSI surveillance systems based on a variety of algorithms using different features. Algorithm performance was disparate, with sensitivities ranging from 20% to 100% and specificities from 59% to 100%. Our model has been designed to minimize false negatives, with performance metrics that are situated in the upper range of the models included in the review. The combination of NLP and XGBoost using data from multiple sources confers a PPV of 19%, compared to 11% for a model using only diagnostic coding^16^, with a better sensitivity (99·7%) than models using NLP alone (94%)^10^ or NLP and logistic regression (97%)^11^, making it the ideal tool for surveillance workload reduction.

The different features included in the model have been evaluated in predictive models for other types of surgical site infection^3,17,18^, with microbiological variables being the most frequent, a finding which is corroborated by the relative importance of positive cultures in our study. NLP-based methods for SSI surveillance have been validated in various pilot studies, showing more accurate results than automated surveillance techniques that rely solely on administrative data such as coding^10^. While diagnostic coding has been included as a feature in multiple studies^19–21^, we decided to exclude it in our model because of the temporal variations between different versions of the International Statistical Classification of Diseases and Related Health Problems (ICD), which could hinder algorithm development. Also, the time lag between patient discharge and diagnostic codification prevents the immediate inclusion of coding variables in the dataset, thus posing an obstacle for real-time surveillance.

Our study has a few limitations. Firstly, the retrospective detection of SSI by infection preventionists at different moments in time could lead to classification error. However, staff expertise and the existence of a standard definition for SSI make this risk negligible. Secondly, our model’s positive predictive value does not permit confirmation of surgical site infection on the sole basis of a positive result. As our primary objective was to reduce the proportion of medical charts to be screened manually, we did not expect to rely on the algorithm to validate positive cases, but instead chose a semi-automatic approach to SSI surveillance, as in many other studies^3,18^. Thirdly, at present, our model can only be used for SSI detection in patients undergoing hip replacement. Although the unbalanced nature of our sample required the use of weights to ensure a high negative predictive value, the overall prevalence of hip prosthetic joint infection in the general population (1·3% to 2·2%)^22^ is similar to that of our sample, indicating that our model could be used successfully in other centers, although further validation is needed. At present, the AI-HPRO SSI surveillance algorithm is fully operative at the four hospitals belonging to the Quironsalud 4-H Network, where it has reduced surveillance workload for hip replacement surgery by over 85%. Next steps include the incorporation of a real-time alert system when the algorithm detects a possible case of SSI, and further algorithm development for other infection control scenarios.

We conclude that an algorithm for SSI surveillance after hip replacement based on NLP techniques and XGBoost is a sensitive tool with a high negative predictive value, permitting real-time, semi-automatic SSI surveillance.

## Supporting information

Members of the Surgical Site Infection Surveillance Group

## Data Availability

All data produced in the present study are available upon reasonable request to the authors.

## Acknowledgements

We would like to thank the UICO (Clinical and Organizational Innovation Unit) for their support of this project. We acknowledge Dr. Bernadette Pfang, M.D. for her help with the writing and editing of the manuscript.

## Notes

### Competing Interest Statement

The authors have declared no competing interest.

### Author Declarations

The ethics committee of the Fundación Jiménez Díaz University Hospital gave ethical approval for this work.

