## Supplementary material for "Design and Retrospective Cohort Validation of an Algorithm for Surgical Site Infection Surveillance after Hip Replacement Surgery Using Natural Language Processing and Extreme Gradient Boosting": Members of the Surgical Site Infection Surveillance Group

| Name | Surname | Role/Highest Degree | Institution |
| --- | --- | --- | --- |
| Gonzalo | De las Casas Cámara | Doctor/Ph.D | Rey Juan Carlos University Hospital |
| Beatriz | Vila Cordero | Doctor/M.D. | Rey Juan Carlos University Hospital |
| Nuria | Gálvez Carranza | Registered Nurse | Rey Juan Carlos University Hospital |
| Carolina | Lucas Molina | Registered Nurse | Fundación Jiménez Díaz University Hospital |
| María Isabel | López Córdoba | Registered Nurse | Infanta Elena University Hospital |
| Marina | Salazar Calzado | Registered Nurse | General Villalba University Hospital |
